# Emergency medical service personnel’s out-of-hospital cardiac arrest case volume and patient outcomes

**DOI:** 10.1101/2025.06.23.25323355

**Authors:** Carsten Meilandt, Lars W. Andersen, Mikael Fink Vallentin

**Affiliations:** Prehospital Emergency Medical Services, Central Denmark Region, Aarhus, Denmark

## Abstract

This document describes the protocol for the study *“Emergency Medical Service Personnel’s Out-of-Hospital Cardiac Arrest Case Volume and Patient Outcomes.”* This will be an observational cohort study using prospectively collected data from the Danish Cardiac Arrest Registry and the Emergency Medical Coordination Centres in Denmark. The exposure will be defined as the number of out-of-hospital cardiac arrest cases attended by each emergency medical service personnel in the preceding year. The primary outcome will be 30-day survival. Generalized linear model with generalized estimating equations will be used to account for correlation within emergency medical service personnel and units.

## Introduction

Out-of-hospital cardiac arrest (OHCA) is a major public health concern worldwide, impacting millions, including approximately 5.000 cases annually in Denmark.^1–4^ Survival is dismal with a 30-day survival rate of 14% in Denmark.^3^ Effective treatment by bystanders and emergency medical services (EMS), including early recognition, high-quality cardiopulmonary resuscitation (CPR), defibrillation, and timely medication, are crucial for improving outcomes.^5–8^

There is some evidence that resuscitation skills decay over time, potentially leading to OHCA treatments falling below the recommended guideline standards.^9^ The number of OHCA cases EMS personnel have been recently exposed to may be a key factor, thus higher OHCA case volume may enhance patient survival.^10–12^ A 2020 systematic review from the International Liaison Committee on Resuscitation suggested that higher exposure to cardiac arrest cases is associated with improved patient outcomes, although this was based on evidence with very low certainty.^13^ Thus, the International Liaison Committee on Resuscitation highlighted the need for further research exploring the relationship between EMS personnel’s OHCA case volume and patient outcomes.^13^

This study aims to examine the association between EMS personnel exposure to OHCA resuscitation and patient outcomes.

### Main aim and hypothesis

The study’s main aim is to estimate the effect of EMS personnel’s OHCA case volume, defined as number of OHCA cases with CPR and/or defibrillation in the preceding year, on patients’ 30-day survival rate. A secondary aim of the study is to assess whether a threshold can be identified for the effect of OHCA case volume on 30-day survival.

The main hypothesis is that 30-day survival increase with increasing EMS personnel OHCA case volume.

## Method

### Study design

This is an observational cohort study based on prospectively collected data from national Danish registries.

### Data source

OHCA data will be obtained from the Danish Cardiac Arrest Registry^14^, where each case is validated by health care professionals. Information regarding the attending EMS personnel will be obtained from the five regional Emergency Medical Coordination Centres and the Danish Air Ambulance database. Patient comorbidities and survival status will be obtained from The National Patient Registry and the Danish Civil Registration System.^15^ EMS personnel information (e.g., level of education) will be requested from local employee databases. To ensure accurate case volume data for EMS personnel, we will apply for employee full name, birth date, and authorisation ID, allowing us to identify EMS personnel’s OHCA case volume across regional boundaries. EMS personnel data will be aggregated regionally, replaced with national authorisation IDs when possible, and combined across regions. To ensure full anonymization of EMS personnel, random identifiers will replace EMS personnel information before linking with patient outcome data to analyse the relationship between OHCA case volume and survival.

Denmark is divided into five regions, each operating its own IT system for dispatching EMS units. Data is expected to be available for four regions. Some regions changed IT systems during the study period, so available data on EMS personnel from the regional Emergency Medical Coordination Centres varies by system and time. The expected data availability is outlined in Table 1.

**Table 1:**
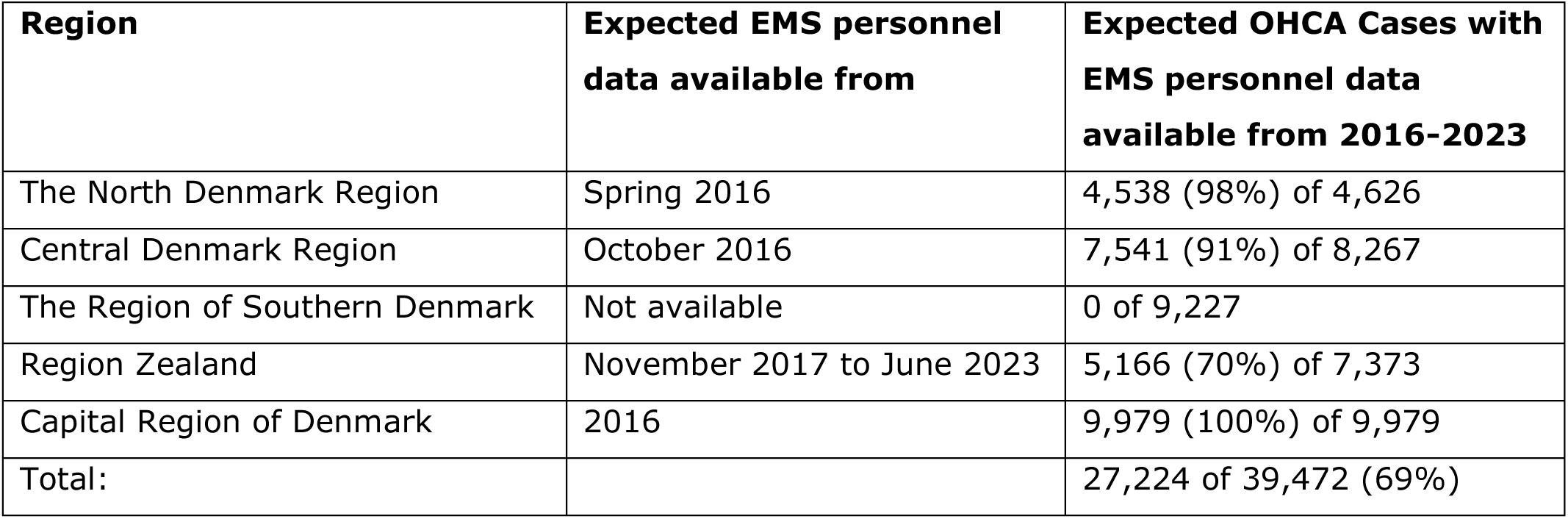
Adult (≥18 years of age) out-of-hospital cardiac arrest cases in Denmark in 2016-2023.

| **Region** | **Expected EMS personnel data available from** | **Expected OHCA Cases with EMS personnel data available from 2016-2023** |
| --- | --- | --- |
| The North Denmark Region | Spring 2016 | 4,538 (98%) of 4,626 |
| Central Denmark Region | October 2016 | 7,541 (91%) of 8,267 |
| The Region of Southern Denmark | Not available | 0 of 9,227 |
| Region Zealand | November 2017 to June 2023 | 5,166 (70%) of 7,373 |
| Capital Region of Denmark | 2016 | 9,979 (100%) of 9,979 |
| Total: |  | 27,224 of 39,472 (69%) |

In our study, EMS personnel exposure will be measured from the first OHCA case with available personnel data (see Table 1) in each region to December 31, 2023. Outcomes for the primary analysis will be measured from one year after the first OHCA case with available personnel data to January 31, 2024 (e.g. in Central Denmark Region, OHCA case volume will be calculated from October 2016 while outcome data will be analysed from October 2017 onwards). This may result in an underestimation of case volume for some EMS personnel (e.g. employed in the Central Denmark Region until September 2016 and subsequently moving to the Capital Region of Denmark, as we cannot retrieve their case volume from the earlier period in the Central Denmark Region).

### Setting

Denmark is divided into five regions responsible for the Danish healthcare system, including the prehospital service and hospitals, and the Danish healthcare system is publicly funded.^16^ In general, Denmark operates a two-tier EMS system.^17^ The first tier consists of an ambulance with either emergency medical technicians or paramedics who primarily provide basic life support, including manual rhythm analysis and defibrillation. Further, the Emergency Medical Coordination Centres can activate volunteer responders through a smart-phone application, which may allow paramedics in the first tier to initiate advanced life support.^18^ The second tier consists of ground based advanced life support units usually operated by a paramedic or an anaesthesiologist and helicopter-based critical care teams consisting of an anaesthesiologist, a pilot, and a specially-trained paramedic.

### Competencies of care

In Denmark, generally three competency levels of prehospital personnel exist.^19^ Emergency medical technician, paramedic, and anaesthesiologist. The emergency medical technician undergoes a 4-year education, including internships in ambulance services and hospitals, and work as trainees under supervision before qualifying. The emergency medical technician provides basic life support and manual defibrillation. Following three years of practice, the emergency medical technician may receive a further five weeks of theoretical and practical education as a paramedic. The paramedics deliver advanced life support. Lastly, the physician-manned rapid response units and the helicopter emergency medical services are manned with consultant anaesthesiologists.

### Study population

For the analyses concerning patient outcomes, we will include all cases of OHCA in patients aged 18 years or older, registered in the Danish Cardiac Arrest Registry and where data on EMS personnel’s OHCA case volume from the regional Emergency Medical Coordination Centres is available for at least one year prior to the OHCA incident. Patients are only entered into the Danish Cardiac Arrest Registry if the patient was defibrillated or EMS personnel initiated CPR.^14^ In cases of bystander CPR without bystander defibrillation, patients are not included in the registry if the EMS personnel does not continue CPR due to overt clinical signs of irreversible death or if the patient shows signs of life. Patients with non-index events and missing data on the primary outcome will be excluded from all outcome analyses. For the primary analysis, patients with any missing or inconsistent data on patient demographics and OHCA characteristics will be excluded. Missing and inconsistent data are expected to some extent, thus a sensitivity analysis using multiple imputation will be performed, with all available variables including the exposure and the outcomes. We expect to have available data from 2016 through 2023 of approximately 27.000 OHCAs.

### Exposure

Exposure to OHCA will be defined as attendance at an OHCA case included in the Danish Cardiac Arrest Registry, where the EMS commenced or continued CPR. This includes cases where CPR was attempted but halted early due to recognition of futility, as discussed later. To measure previous exposure, the OHCA case volume for the individual EMS personnel will be counted in the preceding year before each case.

For our primary analysis, the exposure will be the number of attended OHCA cases in the preceding year for each EMS personnel in the first arriving prehospital unit.

### Outcomes

The primary outcome will be 30-day survival. This will be obtained through the Danish Civil Registration System. Secondary outcomes include any return of spontaneous circulation and patient status at hospital arrival, which will be obtained through the Danish Cardiac Arrest Registry.

### Additional data

Additional variables not mentioned above that will be obtained are listed below.

Required data elements:

**Table 2.**
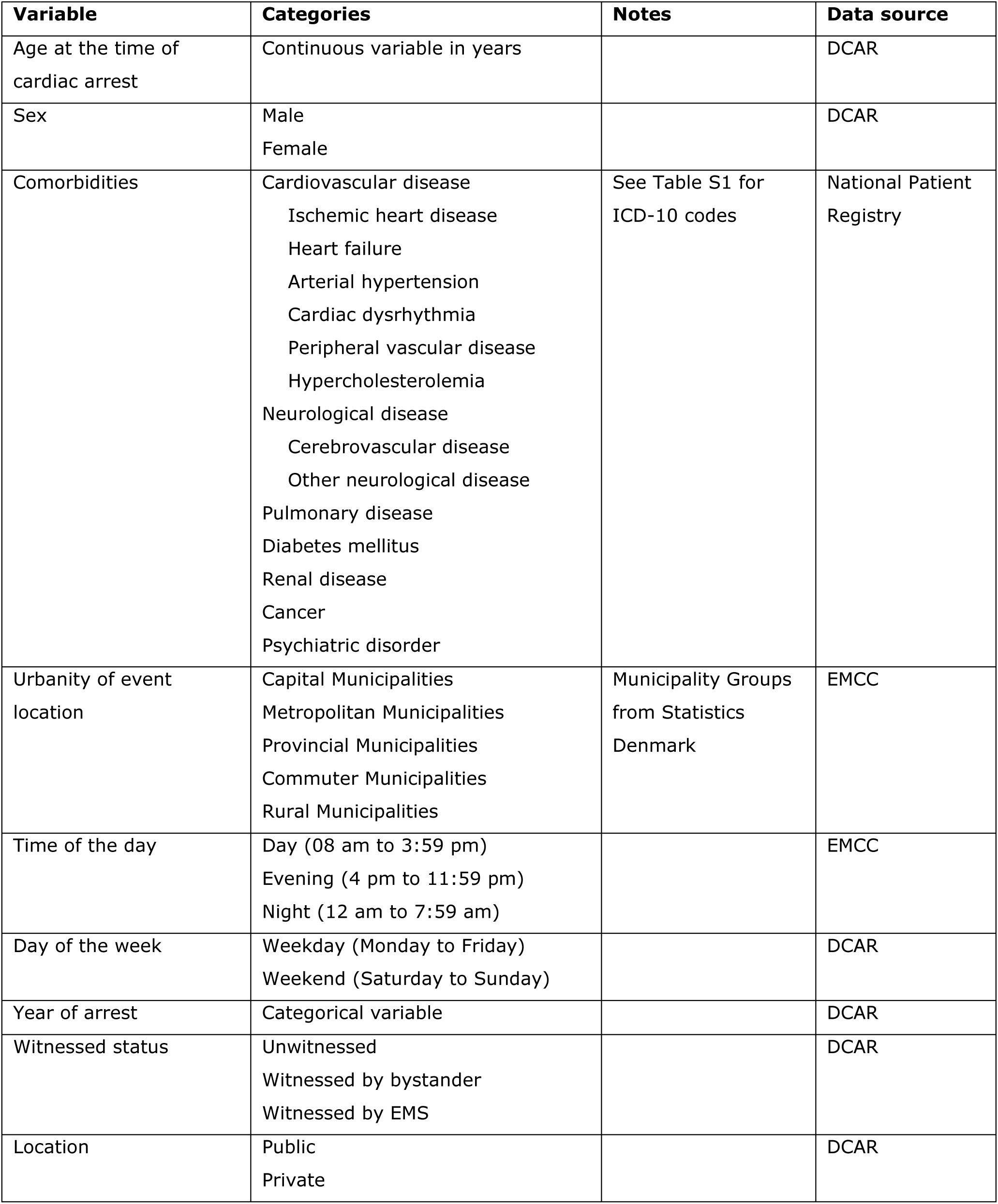
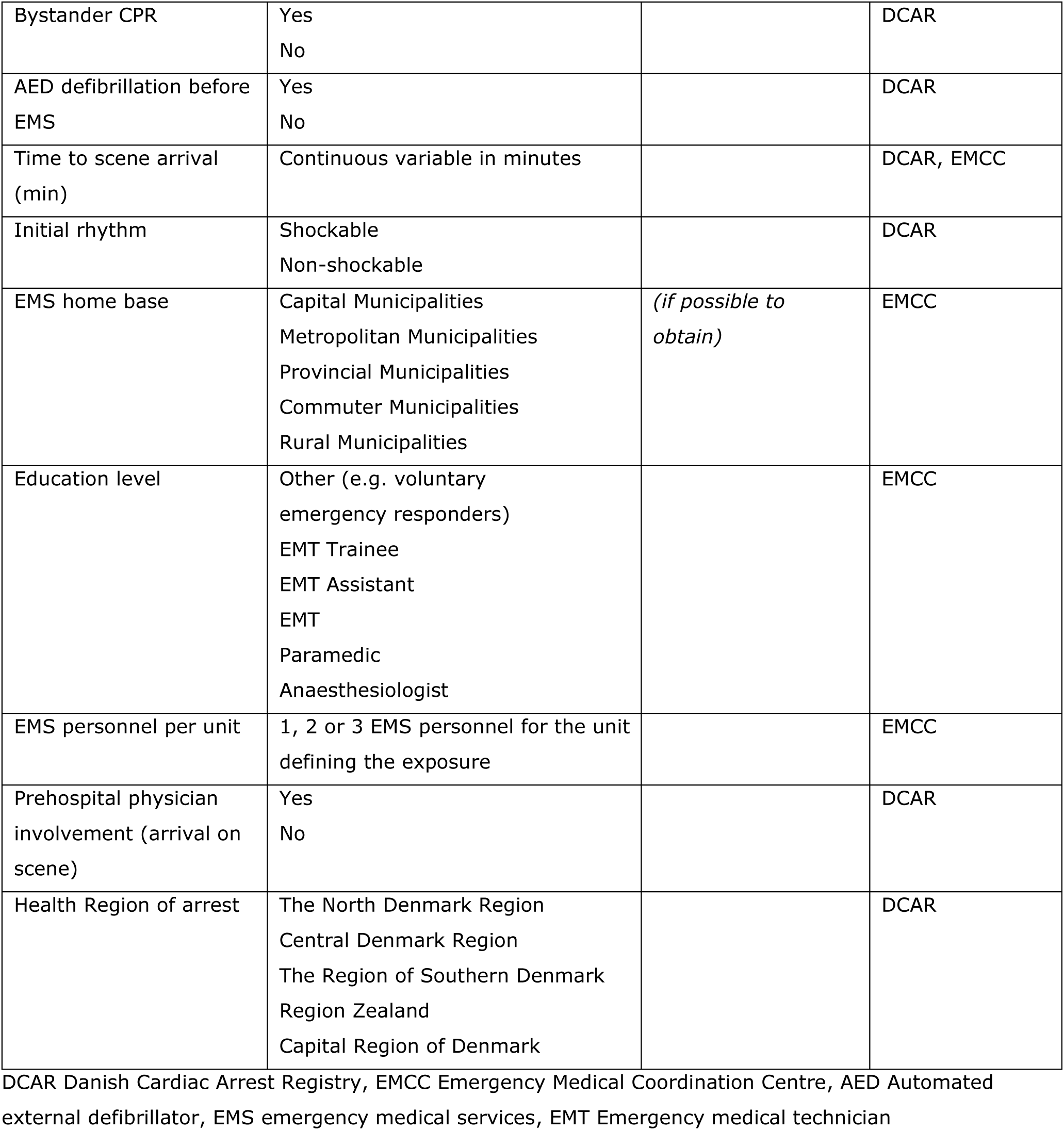

### Statistical analyses

Descriptive data will be presented as numbers (with percentages) for categorical data and means (with standard deviations) for normally distributed continuous data, or medians (with interquartile ranges) for continuous data that is not normally distributed. Both unadjusted and adjusted risk ratios will be reported with 95% confidence interval,^20^ and a two-sided p-value of <0.05 will be considered statistically significant.

In the primary analysis, to examine the association between EMS personnel’s OHCA case volume and patient survival, a multivariable generalized linear model will be used. To account for correlation within EMS units and within EMS personnel, we will implement an extension of the generalized estimating equations as described by Miglioretti & Heagerty.^21^ Risk ratios will be estimated with a binomial distribution and log link function.^22^ If this model fails to converge, a modified Poisson regression model will be used instead (with Poisson distribution and log link function with robust standard errors).^22,23^ First, an unadjusted analysis will be performed. Second, we will adjust for known potential confounders of the exposure-outcome relationship and predictors of survival.^24^ This includes patient demographics and risk factors (age, sex, and comorbidities), urbanity of EMS personnel’s base location (if unable to obtain, a surrogate through the address of the arrest will be used), OHCA factors not potentially mediated by OHCA case volume (number of EMS personnel in the first arriving unit as well as (if able to obtain) EMS personnel education level; (emergency medical technician assistant, emergency medical technician trainee, emergency medical technician, paramedic, or anaesthesiologist), health region, public or private location of arrest, time of the day, day of the week and year of cardiac arrest) and strong predictors of survival (witnessed status, bystander CPR and bystander defibrillation before EMS arrival). To avoid any risk of collider bias, we will not adjust for potential mediators, i.e., initial rhythm (more exposed EMS personnel may be better at identifying a shockable rhythm)^10^ and EMS response time. Age will be included as a continuous variable with linear and quadratic terms,^25^ while the remaining variables will be included as categorical variables. We used directed acyclic graphs to identify relevant covariates for adjustment (Figure 2).

For the primary, adjusted model, polynomial terms for OHCA case volume will be fitted to assess non-linearity between EMS personnel’s OHCA case volume and 30-day survival. OHCA case volume will be included with linear, quadratic, and cubic transformations, and results will be presented graphically with predicted probabilities for primary and secondary outcomes. The Quasi-Likelihood under Independence Model Criterion and visual inspection will be used to evaluate and compare the introduced polynomial terms in the model. If relevant, splines with an appropriate number of knots will be used, based on the best fit model. A potential plateau effect will be identified by visualizing OHCA case volume against predicted probabilities from the best-fitting model.

### Subgroup analysis

We will include a subgroup analysis using our primary model based on whether the OHCA event was EMS-witnessed by the first arriving unit. In cases where EMS witnesses the OHCA, the situation requires an immediate response with no time to discuss or plan. In contrast, when EMS is dispatched to an OHCA, there is time during the drive to discuss and prepare for the actions needed upon arrival. This difference in pre-arrival dynamics may influence how EMS personnel respond according to their previous OHCA case volume.

### Secondary analysis

Although arriving at a later stage during OHCA, the subsequent units may be specialized critical care teams and often with greater OHCA case volume. To assess the association of OHCA case volume and patient survival for EMS personnel arriving as the second unit, the exposure will be the case volume for EMS personnel in the second arriving unit. To account for treatment provided by the first arriving unit, the mean OHCA case volume of EMS personnel in the first arriving unit will be included as covariates in the model.

### Sensitivity analysis

We will conduct several sensitivity analyses:

1) Use 6, 18 and 24-month exposure periods instead of the one-year period.
2) Categorising OHCA case volume in clinical meaningful groups (e.g. 0-2, 3-5, 6-8, 9+ cases in the preceding year with the largest group size as the reference group)
3) Use the mean and max of attended OHCA cases in the preceding year for the EMS personnel in the first arriving prehospital unit. This model will not use generalised estimating equations with clustering but will be adjusted for same covariates except for highest EMS personnel education level on first arriving unit (emergency medical technician, paramedic, or anaesthesiologist).
4) To minimize underestimating OHCA case volume for EMS personnel who may have worked across different regions, we will limit the study period to only include the timeframe where EMS personnel data are available for all the included regions (i.e. from November 10, 2017 to June 11, 2023)
5) Use multiple imputation to account for missing data in our primary analysis. Missing data will be imputed using multiple imputation by chained equations 20 times.^28^ The imputation model will include all variables included in the primary analysis, including exposure and outcome.

### Selection bias

The selection of the patient population and the exposure variable may introduce bias in form of collider bias. Collider bias occurs when an exposure and outcome (or factors causing these) each influence a common third variable (the collider), and that collider is controlled for by design or in the analysis.^26^

In our study, collider bias could be introduced for two reasons (conditioning on attempted resuscitation and by using the first arriving unit as primary exposure). While avoiding risk of collider bias is ideal, we consider these issues of limited importance for the following reasons:

- <u>We only include patients with attempted resuscitation by the EMS.</u>

a. We are conditioning on a variable that could be influenced by both the exposure and factors causing the outcome (Figure 1). EMS personnel with higher OHCA case volumes might be more skilled in assessing which patients have a realistic chance of survival to 30-day. Experienced EMS personnel, based on their case volume, may decide not to initiate resuscitation for patients who could have gained return of spontaneous circulation with attempted resuscitation, albeit potentially with poor neurological outcomes after 30 days. An Australian study by Dyson et al. investigated the associations between OHCA case volume and survival to hospital discharge, as well as OHCA case volume and the odds of attempted resuscitation.^10^ They found that higher OHCA case volume among paramedics was associated with increased patient survival to hospital discharge while highly exposed paramedics (>17 OHCA cases in three years) and longer years of experience (>8 years) was associated with lower odds of attempted resuscitation. However, the association between OHCA case volume and survival to hospital discharge were largely unchanged when comparing OHCA with attempted resuscitation to all OHCA regardless of resuscitation attempt, suggesting a weak association between highly exposed paramedics not attempting resuscitation and 30-day survival. Although we cannot perform a similar sensitivity analysis due to a lack of data on patients without attempted resuscitation, we believe the risk of collider bias is minimal. By adjusting for relevant OHCA factors (e.g., bystander CPR, age, and sex as illustrated in Figure 1), and given Dyson et al.’s similar findings when conditioning on attempted resuscitation, we aim to mitigate potential collider bias from this issue.
b. In Denmark, only physicians are allowed to declare a patient dead, unless the patient is obviously deceased at the non-physician EMS personnel’s arrival (e.g. fatal injuries or trauma that are incompatible with continued life, such as decomposition, charring, or crushing of the skull). As the first arriving units in Denmark primarily are manned with non-physician EMS personnel, obligated to attempt resuscitation (unless a Do Not Attempt Resuscitation order is in place), some patients will be declared dead shortly after EMS physician arrival. We are unable to collect data on how frequently this occurs. However, we believe these cases should still be included for two reasons: i) in relation to the exposure (OHCA case volume): These cases may represent a relevant exposure for the first prehospital personnel, as they are actively involved in managing an OHCA case (e.g., retrieving equipment, initiating CPR, attaching a defibrillator, conducting rhythm analysis, etc.) and ii) in relation to the outcome: Excluding these cases from the analysis would be based on an event that occurs after the initial exposure (i.e., the EMS physician declaring the patient dead), therefore introducing an extra risk of collider bias.
- <u>In our primary analysis, the exposure variable is based on the first arriving unit, excluding other dispatched units (including those that never arrived on scene).</u>

Arguably, once a prehospital unit is dispatched, they could theoretically influence patient outcomes based on their previous exposure (OHCA case volume). For instance, more exposed EMS personnel may better acknowledge the severity of an OHCA case and, in response, drive faster under blue lights, potentially reducing EMS response time. However, we consider it highly unlikely that exposure (OHCA case volume) would significantly affect EMS response time or the decision not to arrive at the scene. The decision not to arrive on scene is made by the Emergency Medical Communication Centre, which has no knowledge of their OHCA case volumes, and this decision is only made when a closer prehospital unit becomes available and is thus not associated with the exposure. We assume that EMS personnel cannot impact patient outcomes until they have made direct contact with the patient. Therefore, we believe it is highly unlikely that this would introduce collider bias. To support this, we will include a supplemental figure with a visual representation of the average EMS response time for the first arriving units, stratified by OHCA case volume.

**Figure 1.**
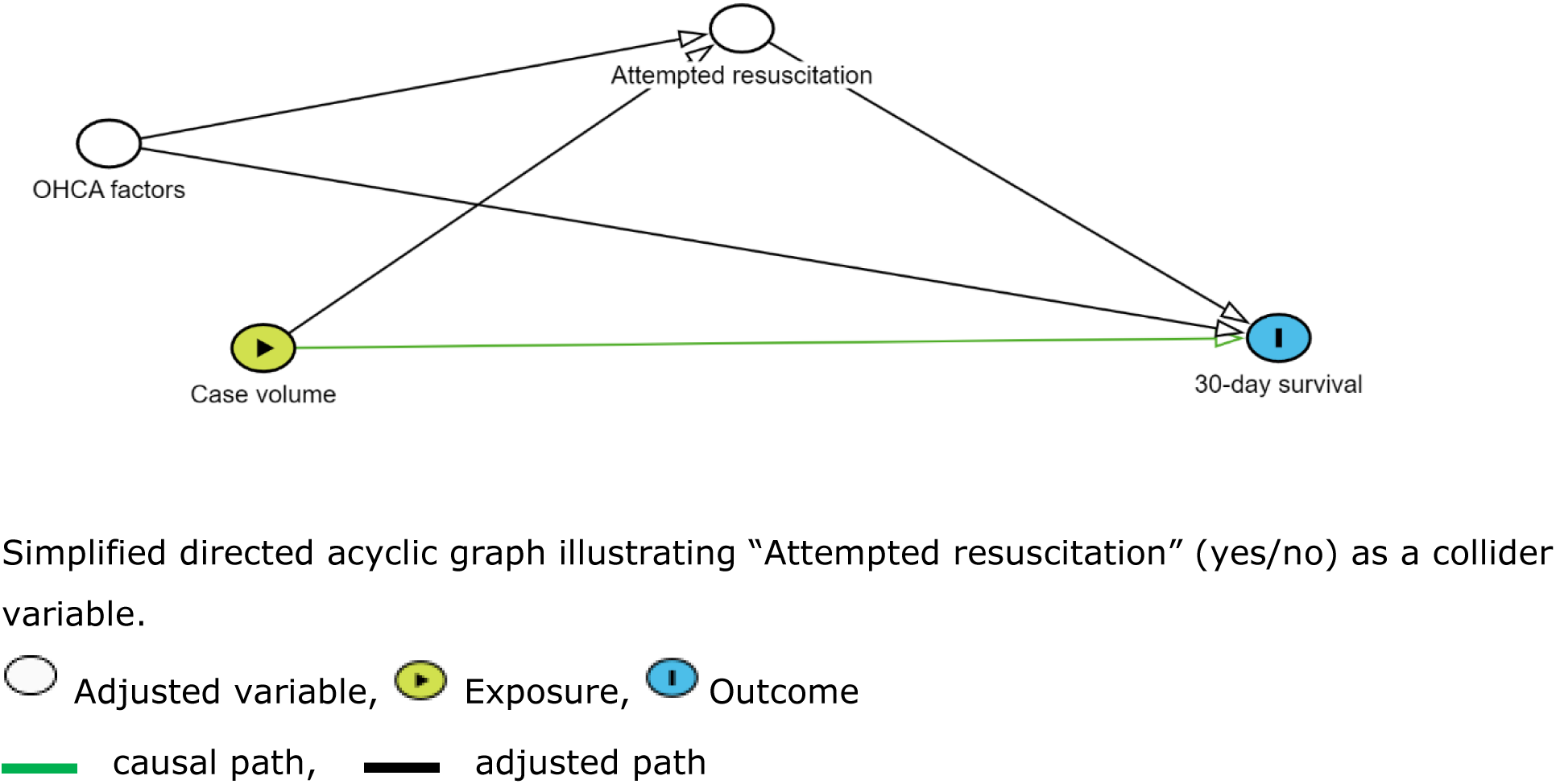
Collider bias - Directed acyclic graph.

**Figure 2.**
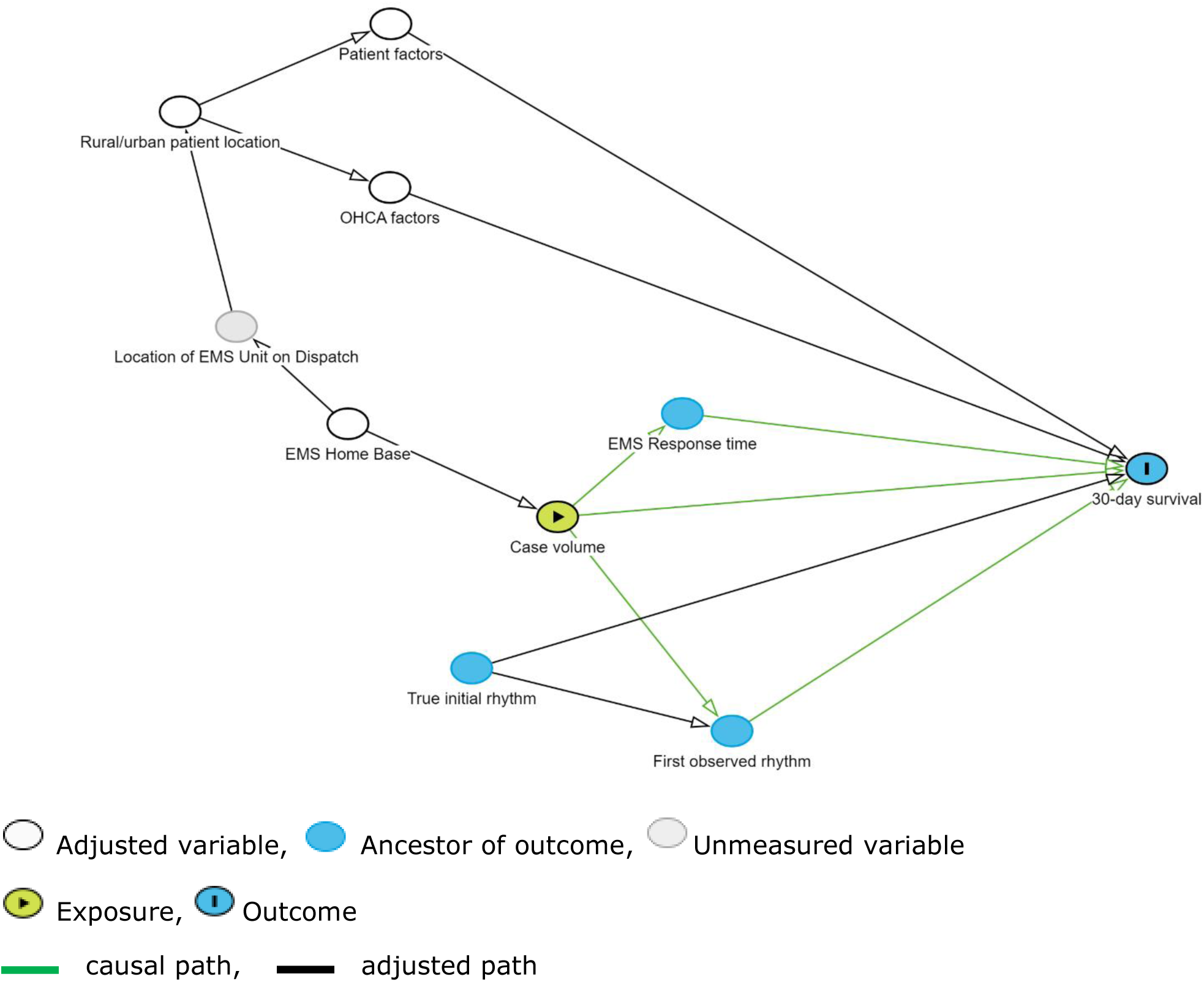
Directed acyclic graph.

### Missing data

We expect missing data on some variables.

Firstly, the regional Emergency Medical Coordination Centres may have transitioned to new computer systems, resulting in an inability to retrieve the requested data regarding which EMS personnel was present at each OHCA case from older or newer systems (Table 1). Such cases are, in any case, excluded from the analyses and this will be assumed to be “missing completely at random”.^27^

Second, some patients treated by EMS may not have a known personal identification number, making it impossible to associate them with comorbidity data. In such cases, these patients will be excluded from the outcome analysis. However, if possible to link the case with the attending EMS personnel’s information, the incident will still be counted towards the total OHCA case volume for those personnel.

Third, missing data for other variables included in the analysis models, including the outcomes, will be missing due to lack of relevant data in the medical records. The overall pattern of missingness is assumed to be “missing at random”.^27^ Missing data will be reported for each variable and a comparison of those with any missing data and those with no missing data will be provided in a supplemental table. As a sensitivity analysis, missing patient data will be imputed using multiple imputation by chained equations 20 times.^28^ The imputation model will include all variables included in the primary analysis.

### Ethical approval

Approval from The Committee on Health Research Ethics and informed consent is not required according to Danish legislation, as this is an observational registry-based study. The study is registered by the data responsible unit in the Central Denmark Region (1-16-02-90-24). The study has received approval for the disclosure of patient journal data (1-45-70-40-24). The study protocol will be published online before any analyses with outcome data.

### Perspectives

The impact of higher EMS personnel exposure to resuscitation cases on patient survival in Denmark is uncertain. If the research shows a positive association with a plateau, implementing geographical rotations, where EMS personnel would rotate from low-exposure to high-exposure areas could potentially enhance overall survival. While promising, careful consideration would need to be given to the logistics, resource availability, and potential push-back from personnel. Furthermore, this research could be advocating for structured and personal cardiac arrest-scenario training. This could be individually tailored based on each EMS personnel’s OHCA case volume, allowing for data-driven training programs and personalized development.

In conclusion, the research may influence EMS practices, training, and resource allocation, ultimately improving patient outcomes in out-of-hospital cardiac arrest.

### Timeline

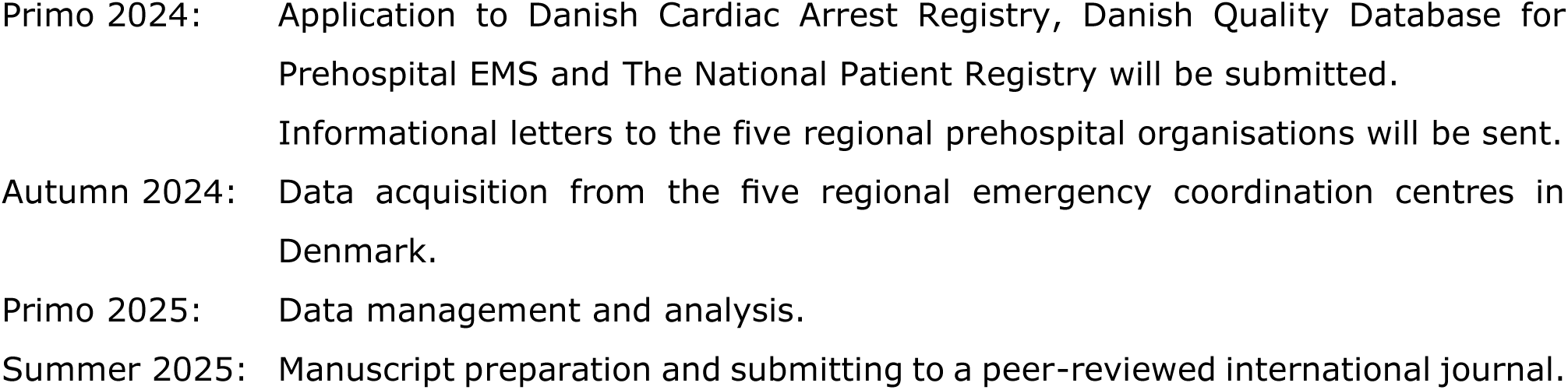

### Investigators

Carsten Meilandt, BSc, Paramedic, Master’s Degree student in Health Science at Aarhus University, is the lead investigator of the study. Associate professor Mikael Fink Vallentin, MD, PhD and professor Lars W. Andersen, MD, MPH, PhD, DMSc have committed themselves as senior researchers and scientific mentors. The lead investigator is responsible for the data, analyses, and preparation of the main paper.

### Data Management

Data will be stored at the Central Denmark Region’s MidtX-site. MidtX is a digital collaboration platform, developed for secure storing of data from research and quality projects. MidtX is thus approved for storage of person-sensitive data and can only be accessed by invitation from the administrator. No data will be disclosed to third parties, and all data will be destroyed at the end of the project.

### Dissemination

The data obtained will only be used for statistical and scientific purposes. Study results will be published irrespective of the findings. The results will be presented at both national and international conferences and the manuscript published in an international peer-reviewed journal. The manuscript will adhere to STROBE reporting guidelines.^29^

## Data Availability

N/A

## Supplemental Tables

**Table S1:** ICD-10 Codes.

| Table S1: ICD-10 Codes |  |  |  |
| --- | --- | --- | --- |
| Central nervous system disease |  |  | References |
| Cerebrovascular disease | DI60<br>DI61<br>DI62<br>DI63<br>DI64<br>DI65<br>DI66<br>DI67<br>DI68<br>DI69<br>DG45<br>DG46 | Subarachnoid haemorrhage<br>Intracerebral haemorrhage<br>Other nontraumatic intracranial haemorrhage<br>Cerebral infarction<br>Stroke, not specified as haemorrhage or infarction<br>Occlusion and stenosis of precerebral arteries, not resulting in cerebral infarction<br>Occlusion and stenosis of cerebral arteries, not resulting in cerebral infarction<br>Other cerebrovascular diseases<br>Cerebrovascular disorders in diseases classified elsewhere<br>Sequelae of cerebrovascular disease<br>Transient cerebral ischaemic attacks and related syndromes<br>Vascular syndromes of brain in cerebrovascular diseases | (1-3) |
| Other neurological disease | DG20<br>DG21<br>DG22<br>DG81<br>DG82<br>DF00<br>DF01<br>DF02<br>DF03<br>DF051<br>DG30<br>DG40<br>DG41 | Parkinson disease<br>Secondary parkinsonism<br>Parkinsonism in diseases classified elsewhere<br>Hemiplegia<br>Paraplegia and tetraplegia<br>Dementia in Alzheimer disease<br>Vascular dementia<br>Dementia in other diseases classified elsewhere<br>Unspecified dementia<br>Delirium superimposed on dementia<br>Alzheimer disease<br>Epilepsy<br>Status epilepticus | (1, 4-6) |
| Cardiovascular disease |  |  |  |
| Ischemic heart disease | DI20<br>DI21<br>DI23<br>DI24<br>DI25<br>DT823D<br>DT823E | Angina pectoris<br>Acute myocardial infarction<br>Certain current complications following acute myocardial infarction<br>Other acute ischaemic heart diseases<br>Chronic ischaemic heart disease<br>Acute coronary stent thrombosis<br>Coronary stent restenosis | (7) |
| Heart failure | DI110<br>DI130<br>DI132<br>DI420<br>DI426<br>DI427<br>DI428<br>DI429<br>DI500<br>DI501<br>DI509 | Hypertensive heart disease with (congestive) heart failure<br>Hypertensive heart and renal disease with (congestive) heart failure<br>Hypertensive heart and renal disease with both (congestive) heart failure and renal failure<br>Dilated cardiomyopathy<br>Alcoholic cardiomyopathy<br>Cardiomyopathy due to drugs and other external agents<br>Other cardiomyopathies<br>Cardiomyopathy, unspecified<br>Congestive heart failure<br>Left ventricular failure<br>Heart failure, unspecified | (7) |
| Arterial hypertension | DI10<br>DI11<br>DI12<br>DI13<br>DI15 | Essential (primary) hypertension<br>Hypertensive heart disease<br>Hypertensive renal disease<br>Hypertensive heart and renal disease<br>Secondary hypertension | (7) |
| Atrial fibrillation or flutter | DI48 | Atrial fibrillation and flutter | (7, 8) |
| Other cardiac dysrhythmias | DI44<br>DI45<br>DI47<br>DI49 | Atrioventricular and left bundle-branch block<br>Other conduction disorders<br>Paroxysmal tachycardia<br>Other cardiac arrhythmias | (9) |
| Peripheral vascular disease | DI70<br>DI72<br>DI73<br>DI77 | Atherosclerosis<br>Other aneurysm and dissection<br>Other peripheral vascular diseases<br>Other disorders of arteries and arterioles | (1) |
| Hypercholesterolemia | DE780 | Pure hypercholesterolaemia | (7) |
| **Pulmonary disease** |  |  |  |
| Chronic obstructive pulmonary disease | DJ40<br>DJ41<br>DJ42<br>DJ43<br>DJ44<br>DJ47 | Bronchitis, not specified as acute or chronic<br>Simple and mucopurulent chronic bronchitis<br>Unspecified chronic bronchitis<br>Emphysema<br>Other chronic obstructive pulmonary disease<br>Bronchiectasis | (10, 11) |
| Asthma | DJ45<br>DJ46 | Asthma<br>Status asthmaticus | (11, 12) |
| **Endocrine disease** |  |  |  |
| Diabetes mellitus | DE10<br>DE11 | Type 1 diabetes mellitus<br>Type 2 diabetes mellitus | (1, 13) |
| **Renal disease** |  |  |  |
| Moderate/severe renal disease | DN00<br>DN01<br>DN02<br>DN03<br>DN04<br>DN05<br>DN07<br>DN11<br>DI12<br>DI13<br>DN14<br>DN17<br>DN18<br>DN19<br>DQ61 | Acute nephritic syndrome<br>Rapidly progressive nephritic syndrome<br>Recurrent and persistent haematuria<br>Chronic nephritic syndrome<br>Nephrotic syndrome<br>Unspecified nephritic syndrome<br>Hereditary nephropathy, not elsewhere classified<br>Chronic tubulo-interstitial nephritis<br>Hypertensive renal disease<br>Hypertensive heart and renal disease<br>Drug- and heavy-metal-induced tubulo-interstitial and tubular conditions<br>Acute renal failure<br>Chronic kidney disease<br>Unspecified kidney failure<br>Cystic kidney disease | (1) |
| **Psychiatric disorder** |  |  |  |
| Psychiatric disease, overall | DF0<br>DF1<br>DF2<br>DF3<br>DF4<br>DF5<br>DF6<br>DF7<br>DF8<br>DF9 | Mental and behavioural disorders [DF00-DF99] | (9) |
| **Cancer** |  |  |  |
| All cancers | DC0<br>DC1<br>DC2<br>DC3<br>DC4<br>DC5<br>DC6<br>DC7<br>DC8<br>DC9 | Neoplasms [DC00-96] | (1, 11, 14-16) |

## References

1. Gräsner JT, Wnent J, Herlitz J, et al. Survival after out-of-hospital cardiac arrest in Europe - Results of the EuReCa TWO study. Resuscitation. 2020;148:218–226. doi:10.1016/j.resuscitation.2019.12.042

2. Berdowski J, Berg RA, Tijssen JGP, Koster RW. Global incidences of out-of-hospital cardiac arrest and survival rates: Systematic review of 67 prospective studies. Resuscitation. 2010;81(11):1479–1487. doi:10.1016/j.resuscitation.2010.08.006

3. Danish Cardiac Arrest Registry. Annual Report 2022. 2022. Accessed November 19, 2023. https://www.sundhed.dk/content/cms/4/123004_dhsr_aarsrapport_2022_endelig.pdf

4. Nishiyama C, Kiguchi T, Okubo M, et al. Three-year trends in out-of-hospital cardiac arrest across the world: Second report from the International Liaison Committee on Resuscitation (ILCOR). Resuscitation. 2023;186:109757. doi:10.1016/j.resuscitation.2023.109757

5. Chan PS, Krumholz HM, Nichol G, et al. Delayed time to defibrillation after in-hospital cardiac arrest. N Engl J Med. 2008;358(1):9–17. doi:10.1056/NEJMoa0706467

6. Deakin CD. The chain of survival: Not all links are equal. Resuscitation. 2018;126:80–82. doi:10.1016/j.resuscitation.2018.02.012

7. Al-Dury N, Ravn-Fischer A, Hollenberg J, et al. Identifying the relative importance of predictors of survival in out of hospital cardiac arrest: a machine learning study. Scand J Trauma Resusc Emerg Med. 2020;28(1):60. doi:10.1186/s13049-020-00742-9

8. Gregers MCT, Andelius L, Kjoelbye JS, et al. Association Between Number of Volunteer Responders and Interventions Before Ambulance Arrival for Cardiac Arrest. J Am Coll Cardiol. 2023;81(7):668–680. doi:10.1016/j.jacc.2022.11.047

9. Yang CW, Yen ZS, McGowan JE, et al. A systematic review of retention of adult advanced life support knowledge and skills in healthcare providers. Resuscitation. 2012;83(9):1055–1060. doi:10.1016/j.resuscitation.2012.02.027

10. Dyson K, Bray JE, Smith K, Bernard S, Straney L, Finn J. Paramedic Exposure to Out-of-Hospital Cardiac Arrest Resuscitation Is Associated With Patient Survival. Circ Cardiovasc Qual Outcomes. 2016;9(2):154–160. doi:10.1161/CIRCOUTCOMES.115.002317

11. Weiss N, Ross E, Cooley C, et al. Does Experience Matter? Paramedic Cardiac Resuscitation Experience Effect on Out-of-Hospital Cardiac Arrest Outcomes. Prehosp Emerg Care. 2018;22(3):332–337. doi:10.1080/10903127.2017.1392665

12. Tuttle JE, Hubble MW. Paramedic Out-of-hospital Cardiac Arrest Case Volume Is a Predictor of Return of Spontaneous Circulation. West J Emerg Med. 2018;19(4):654–659. doi:10.5811/westjem.2018.3.37051

13. Bray J, Nehme Z, Nguyen A, Lockey A, Finn J. A systematic review of the impact of emergency medical service practitioner experience and exposure to out of hospital cardiac arrest on patient outcomes. Resuscitation. 2020;155:134–142. doi:10.1016/j.resuscitation.2020.07.025

14. Jensen TW, Blomberg SN, Folke F, et al. The National Danish Cardiac Arrest Registry for Out-of-Hospital Cardiac Arrest – A Registry in Transformation. Clin Epidemiol. 2022;14:949–957. doi:10.2147/CLEP.S374788

15. Schmidt M, Schmidt SAJ, Sandegaard JL, Ehrenstein V, Pedersen L, Sørensen HT. The Danish National Patient Registry: a review of content, data quality, and research potential. Clin Epidemiol. 2015;7:449–490. doi:10.2147/CLEP.S91125

16. Regional Denmark. Regional tasks in Denmark. Accessed November 24, 2023. https://www.regioner.dk/services/in-english/regional-denmark

17. Lindskou TA, Mikkelsen S, Christensen EF, et al. The Danish prehospital emergency healthcare system and research possibilities. Scand J Trauma Resusc Emerg Med. 2019;27(1):100. doi:10.1186/s13049-019-0676-5

18. Andelius L, Malta Hansen C, Lippert FK, et al. Smartphone Activation of Citizen Responders to Facilitate Defibrillation in Out-of-Hospital Cardiac Arrest. J Am Coll Cardiol. 2020;76(1):43–53. doi:10.1016/j.jacc.2020.04.073

19. Mikkelsen S, Lassen AT. The Danish prehospital system. Eur J Emerg Med. 2020;27(6):394–395. doi:10.1097/MEJ.0000000000000774

20. Holmberg MJ, Andersen LW. Estimating Risk Ratios and Risk Differences: Alternatives to Odds Ratios. JAMA. 2020;324(11):1098. doi:10.1001/jama.2020.12698

21. Miglioretti DL, Heagerty PJ. Marginal Modeling of Nonnested Multilevel Data using Standard Software. Am J Epidemiol. 2006;165(4):453–463. doi:10.1093/aje/kwk020

22. Cummings P. Methods for Estimating Adjusted Risk Ratios. Stata J Promot Commun Stat Stata. 2009;9(2):175–196. doi:10.1177/1536867X0900900201

23. Zou G. A Modified Poisson Regression Approach to Prospective Studies with Binary Data. Am J Epidemiol. 2004;159(7):702–706. doi:10.1093/aje/kwh090

24. VanderWeele TJ. Principles of confounder selection. Eur J Epidemiol. 2019;34(3):211–219. doi:10.1007/s10654-019-00494-6

25. Andersen LW, Bivens MJ, Giberson T, et al. The relationship between age and outcome in out-of-hospital cardiac arrest patients. Resuscitation. 2015;94:49–54. doi:10.1016/j.resuscitation.2015.05.015

26. Holmberg MJ, Andersen LW. Collider Bias. JAMA. 2022;327(13):1282-1283. doi:10.1001/jama.2022.1820

27. Sterne JAC, White IR, Carlin JB, et al. Multiple imputation for missing data in epidemiological and clinical research: potential and pitfalls. BMJ. 2009;338(jun29 1):b2393–b2393. doi:10.1136/bmj.b2393

28. White IR, Royston P, Wood AM. Multiple imputation using chained equations: Issues and guidance for practice. Stat Med. 2011;30(4):377–399. doi:10.1002/sim.4067

29. von Elm E, Altman DG, Egger M, et al. The Strengthening the Reporting of Observational Studies in Epidemiology (STROBE) statement: guidelines for reporting observational studies. Lancet Lond Engl. 2007;370(9596):1453–1457. doi:10.1016/S0140-6736(07)61602-X

## Supplemental References

1. Thygesen SK, Christiansen CF, Christensen S, Lash TL, Sorensen HT. The predictive value of ICD-10 diagnostic coding used to assess Charlson comorbidity index conditions in the population-based Danish National Registry of Patients. BMC Med Res Methodol. 2011;11:83.

2. Krarup LH, Boysen G, Janjua H, Prescott E, Truelsen T. Validity of stroke diagnoses in a National Register of Patients. Neuroepidemiology. 2007;28(3):150–4.

3. Wildenschild C, Mehnert F, Thomsen RW, Iversen HK, Vestergaard K, Ingeman A, et al. Registration of acute stroke: validity in the Danish Stroke Registry and the Danish National Registry of Patients. Clin Epidemiol. 2014;6:27–36.

4. Erlangsen A, Stenager E, Conwell Y, Andersen PK, Hawton K, Benros ME, et al. Association Between Neurological Disorders and Death by Suicide in Denmark. JAMA. 2020;323(5):444–54.

5. Rugbjerg K, Ritz B, Korbo L, Martinussen N, Olsen JH. Risk of Parkinson’s disease after hospital contact for head injury: population based case-control study. BMJ. 2008;337:a2494.

6. Christensen J, Vestergaard M, Olsen J, Sidenius P. Validation of epilepsy diagnoses in the Danish National Hospital Register. Epilepsy Res. 2007;75(2-3):162–70.

7. Sundboll J, Adelborg K, Munch T, Froslev T, Sorensen HT, Botker HE, et al. Positive predictive value of cardiovascular diagnoses in the Danish National Patient Registry: a validation study. BMJ Open. 2016;6(11):e012832.

8. Rix TA, Riahi S, Overvad K, Lundbye-Christensen S, Schmidt EB, Joensen AM. Validity of the diagnoses atrial fibrillation and atrial flutter in a Danish patient registry. Scand Cardiovasc J. 2012;46(3):149–53.

9. Granfeldt A, Adelborg K, Wissenberg M, Moller Hansen S, Torp-Pedersen C, Christensen EF, et al. Severity of ischemic heart disease and presenting rhythm in patients with out-of-hospital cardiac arrest. Resuscitation. 2018;130:174–81.

10. Thomsen RW, Lange P, Hellquist B, Frausing E, Bartels PD, Krog BR, et al. Validity and underrecording of diagnosis of COPD in the Danish National Patient Registry. Respir Med. 2011;105(7):1063–8.

11. Hvidberg MF, Johnsen SP, Glumer C, Petersen KD, Olesen AV, Ehlers L. Catalog of 199 register-based definitions of chronic conditions. Scand J Public Health. 2016;44(5):462–79.

12. Jensen AO, Nielsen GL, Ehrenstein V. Validity of asthma diagnoses in the Danish National Registry of Patients, including an assessment of impact of misclassification on risk estimates in an actual dataset. Clin Epidemiol. 2010;2:67–72.

13. Thomsen RW, Hundborg HH, Lervang HH, Johnsen SP, Sorensen HT, Schonheyder HC. Diabetes and outcome of community-acquired pneumococcal bacteremia: a 10-year population-based cohort study. Diabetes Care. 2004;27(1):70–6.

14. Helqvist L, Erichsen R, Gammelager H, Johansen MB, Sorensen HT. Quality of ICD-10 colorectal cancer diagnosis codes in the Danish National Registry of Patients. Eur J Cancer Care (Engl). 2012;21(6):722–7.

15. Norgaard M, Skriver MV, Gregersen H, Pedersen G, Schonheyder HC, Sorensen HT. The data quality of haematological malignancy ICD-10 diagnoses in a population-based hospital discharge registry. Eur J Cancer Prev. 2005;14(3):201–6.

16. Gammelager H, Christiansen CF, Johansen MB, Borre M, Schoonen M, Sorensen HT. Quality of urological cancer diagnoses in the Danish National Registry of Patients. Eur J Cancer Prev. 2012;21(6):545–51.

